# Efficacy and Safety of Atorvastatin as Adjunctive Treatment in Chronic Obstructive Pulmonary Disease: A Systematic Review and Meta-Analysis

**DOI:** 10.1101/2024.12.04.24318509

**Authors:** Ke Chen, Bowen Xu, Lu Zhang, Li Fang, Di Wu, Huanzhang Ding, Zegeng Li

**Affiliations:** The First Clinical Medical College of Anhui University of Chinese Medicine, Hefei, 230031, China; Anhui University of Chinese Medicine, Hefei,230012, China; Anhui Provincial Chinese Medicine Hospital, Hefei, 230031, China; Institute of Traditional Chinese Medicine for Respiratory Disease Prevention and Treatment, Anhui Academy of Traditional Chinese Medicine, Hefei, 230031; Anhui Provincial Key Laboratory of Translational Medicine for Prevention and Treatment of Major Respiratory Diseases with Traditional Chinese Medicine, Hefei, 230031

**Keywords:** Atorvastatin, chronic obstructive pulmonary, Meta-analysis, RCT

## Abstract

**Background:** Accumulating evidence suggests that atorvastatin, a widely used lipid-lowering agent, may provide additional benefits for chronic obstructive pulmonary disease (COPD) patients, including anti-inflammatory effects and improved lung function. However, inconsistent findings across studies warrant a systematic evaluation to clarify its clinical role.

**Objective:** To systematically evaluate the efficacy and safety of atorvastatin as an adjunctive treatment for COPD and inform clinical decision-making.

**Methods:** A comprehensive search of PubMed, EMBASE, Web of Science, Cochrane Library, CNKI, WanFang, CBM, and VIP databases identified randomized controlled trials (RCTs) up to May 20, 2024. Meta-analysis using RevMan 5.3 and R software was performed to estimate pooled effects with mean differences or standardized mean differences (95% CI). Subgroup analyses explored variations by treatment duration and dosage.

**Results:** Twenty-four RCTs involving 2,534 patients demonstrated significant benefits of atorvastatin for stable COPD and acute exacerbations (AECOPD):Lung function: FEV1%pred increased by 5.36% (95% CI: 4.57–6.14), FEV1/FVC by 6.30% (95% CI: 4.46–8.14), and FEV1 by 0.21 L (95% CI: 0.15–0.27).Inflammatory markers: CRP decreased by 1.87 mg/L (95% CI: 1.45–2.29), with reductions in hs-CRP and IL-6.Quality of life: CAT scores improved by 3.5 points (95% CI: 2.8–4.2).Exercise capacity: The 6-minute walk distance increased by 25.4 meters (95% CI: 18.1– 32.7).Stronger evidence emerged with 3-month treatments (I² < 30%) and consistent benefits at 20 mg doses. Adverse events were mild and self-limiting.

**Conclusion:** Atorvastatin (20 mg) significantly improves lung function, reduces inflammation, and enhances quality of life in COPD patients, with a favorable safety profile. Although not currently recommended in COPD guidelines, these findings support further trials to validate its potential role in COPD management.

## 1. Introduction

Chronic Obstructive Pulmonary Disease (COPD) is a prevalent global public health concern characterized by airflow limitation and chronic airway inflammation[1]. According to data from the World Health Organization, COPD is among the leading causes of mortality globally. Epidemiological studies indicate that the global prevalence of COPD among adults aged 40 and above is approximately 9%∼10%[2].

COPD detrimentally affects patients’ quality of life and imposes a substantial economic burden on healthcare systems[3;4] Although bronchodilators and corticosteroids are commonly utilized in COPD management, their efficacy remains suboptimal in certain patient populations[5]. Recent years have seen the emergence of statins, particularly atorvastatin and simvastatin, as promising agents in the treatment of COPD. Research suggests that atorvastatin may enhance lung function, inhibit inflammatory responses, and reduce oxidative stress in COPD patients while also exhibiting favorable safety profiles[6;7]. Consequently, this study systematically assesses the efficacy and safety of atorvastatin as an adjunctive treatment for COPD, aiming to furnish clinicians with enhanced evidence-based support. Furthermore, it conducts subgroup analyses stratified by dosage, treatment duration, and disease severity, with the goal of providing robust data to inform clinical decision-making.

## 2. Methods

### 2.1. Protocol and Registration

This meta-analysis was conducted in accordance with the Preferred Reporting Items for Systematic Reviews and Meta-Analyses (PRISMA) guidelines(**see Supplementary Document** 1).The protocol for this study was registered in the International Prospective Register of Systematic Reviews (PROSPERO) under registration number CRD42024531466.

### 2.2 Search Strategy and Inclusion Criteria

A comprehensive search of relevant studies was conducted across eight major databases: PubMed, EMBASE, Web of Science, Cochrane Library, CNKI, WanFang Data, CBM, and VIP. The search spanned from the inception of each database to May 20, 2024. To ensure the search captured all relevant studies, we used a combination of Medical Subject Headings (MeSH) and free-text terms, such as “Chronic Obstructive Pulmonary Disease,” “atorvastatin,” “inflammation,” and “lung function.” Boolean operators (AND, OR) were applied, and search terms were tailored to each database. We also conducted supplementary searches, including reference lists of relevant articles and systematic reviews, to identify additional eligible studies.

Studies were eligible for inclusion if they were randomized controlled trials (RCTs) evaluating atorvastatin as an adjunctive treatment in patients diagnosed with chronic obstructive pulmonary disease (COPD) based on established criteria, such as the Global Initiative for Chronic Obstructive Lung Disease (GOLD) guidelines[8]. To ensure clinical relevance, included studies needed to report at least one of the following outcomes: (1) primary outcomes—lung function parameters (e.g., FEV1%pred, FEV1/FVC) and systemic inflammatory markers (e.g., CRP, hs-CRP, IL-6); or (2) secondary outcomes—exercise capacity (e.g., 6-minute walk distance), quality of life (e.g., COPD Assessment Test [CAT]), or adverse events.Interventions were considered eligible if atorvastatin was compared to placebo or standard COPD treatment.

Studies were excluded if they were non-randomized trials, observational studies, case reports, or reviews. Additionally, we excluded studies with insufficient data for analysis, animal models, or those involving atorvastatin doses outside the clinical guidelines. Full-text articles not available in English or Chinese were also excluded to ensure accurate data interpretation.

### 2.3. Data Extraction and Quality Assessment

Data extraction was independently conducted by two reviewers, Ke Chen and Bowen Xu, using a standardized data extraction form to ensure consistency and accuracy. The information extracted included study characteristics, such as author names, publication year, country, and sample size, as well as participant demographics, including age, sex, Chronic Obstructive Pulmonary Disease (COPD) severity, and comorbidities. Details of the interventions, including atorvastatin dosage, treatment duration, and comparator interventions, were also collected. The primary outcomes of interest were lung function parameters, specifically the Forced Expiratory Volume in the first second as a percentage of the predicted value (FEV1%pred) and the Forced Expiratory Volume in the first second to Forced Vital Capacity ratio (FEV1/FVC). Additionally, inflammatory markers, such as C-reactive protein (CRP), high-sensitivity C-reactive protein (hs-CRP), and interleukin-6 (IL-6), were included as primary outcomes. Secondary outcomes encompassed exercise capacity, measured by the six-minute walk distance (6MWD), quality of life, assessed using the COPD Assessment Test (CAT), and adverse events, including elevated alanine aminotransferase (ALT) levels and myopathy. Any disagreements arising during the data extraction process were resolved through discussion or, when necessary, consultation with a third reviewer, Huanzhang Ding, to ensure the reliability of the extracted data.

The quality of included studies was assessed using the Cochrane RoB 2 tool[9], which evaluates five domains: randomization process, deviations from intended interventions, missing outcome data, measurement of outcomes, and selection of reported results. Each domain was rated as “low risk,” “some concerns,” or “high risk,” and overall bias for each study was determined. Two reviewers conducted the risk of bias assessment independently, with discrepancies resolved by consensus. This systematic approach ensured the reliability and validity of the included studies for subsequent meta-analysis.

### 2.4. Data Synthesis and Statistical Analysis

All statistical analyses were performed using RevMan 5.3 and R (v3.6.1). Continuous outcomes (e.g., lung function parameters, inflammatory markers, and quality of life scores) were summarized as mean differences (MD) or standardized mean differences (SMD) with corresponding 95% confidence intervals (CI). Dichotomous outcomes (e.g., adverse events) were analyzed using risk ratios (RR) with 95% CI. The pooled effect size for each outcome was calculated using either a fixed-effect model or a random-effects model, depending on the degree of heterogeneity observed.

Heterogeneity was assessed using the I² statistic and τ² values. An I² > 50% was considered indicative of substantial heterogeneity, prompting the use of a random-effects model to account for variability among studies. For outcomes with low heterogeneity (I² ≤ 50%), a fixed-effect model was employed. Additionally, the Cochran Q test (p < 0.10) was used to further evaluate statistical heterogeneity. Sensitivity analyses were conducted to assess the robustness of the pooled results by excluding high-risk studies (e.g., those rated as “Some Concerns” or “High Risk” in the Cochrane RoB 2 tool) and by re-analyzing data using alternative statistical models.

Subgroup analyses were pre-specified to explore potential sources of heterogeneity. Stratifications were performed based on atorvastatin dosage (20 mg vs. 40 mg), treatment duration (<6 months vs. ≥6 months), and patient condition (stable COPD vs. acute exacerbation of COPD [AECOPD]).

Publication bias was evaluated using funnel plots and Egger’s regression test, provided that the number of included studies exceeded 10, as smaller samples may yield unreliable assessments. Visual inspection of funnel plot symmetry was used to detect potential small-study effects. In cases where asymmetry was observed, additional analyses were conducted to explore its impact on the overall meta-analytic results. Statistical significance was set at p < 0.05 for all analyses, except where otherwise specified.

## 3. Results

### 3.1. Literature Screening Process and Results

A total of 1,100 articles were initially identified. After sequential screening, 24 randomized controlled trials (RCTs)[10; 11; 12; 13; 14; 15; 16; 17; 18; 19; 20; 21; 22; 23; 24; 25; 26; 27; 28; 29; 30; 31; 32; 33],were finally included, involving 2,534 patients with chronic obstructive pulmonary disease (COPD). The literature screening process and results are shown in **Figure 1**.

**Figure 1:**
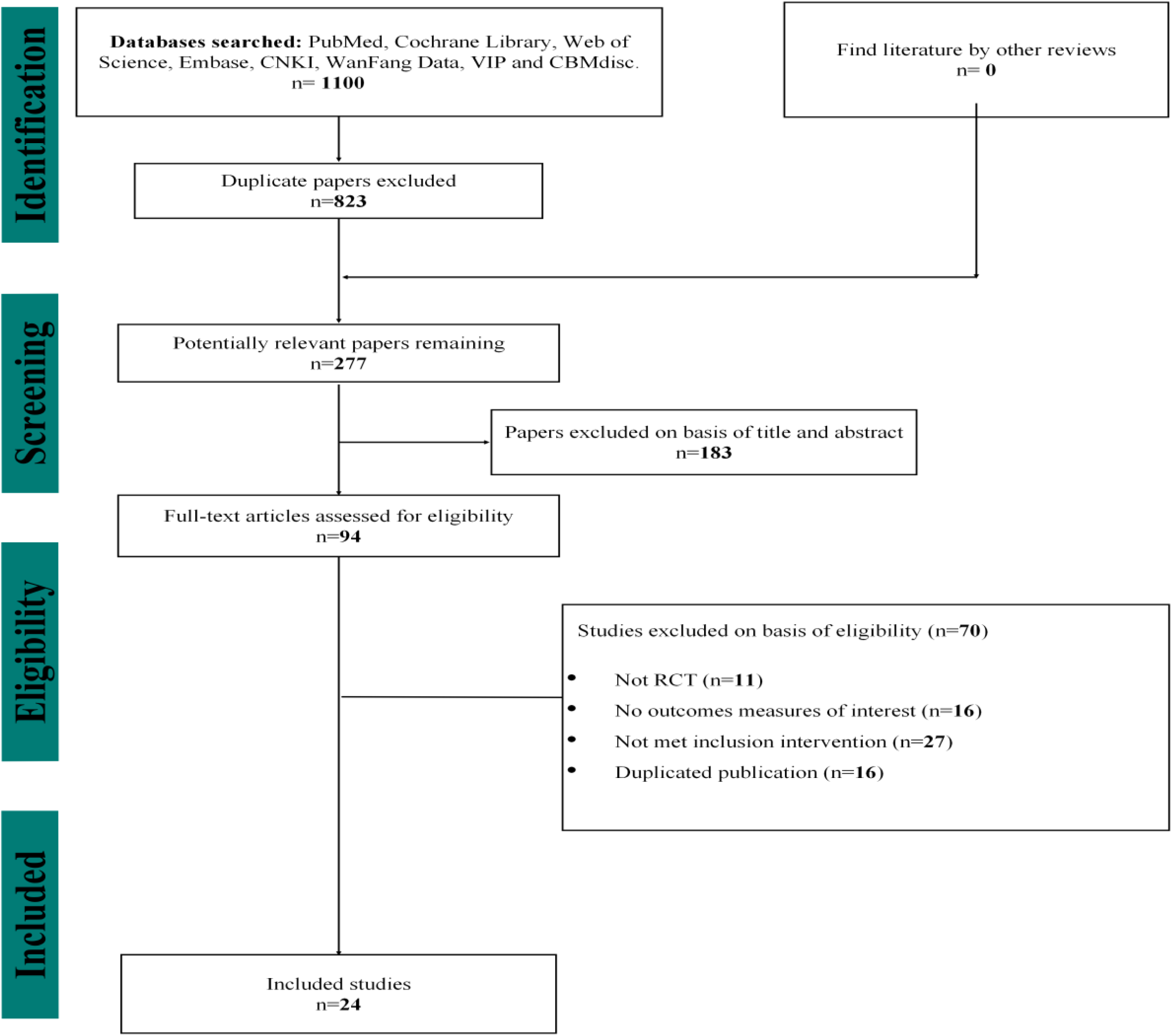
Literature screening process and results.

**Figure 2.**
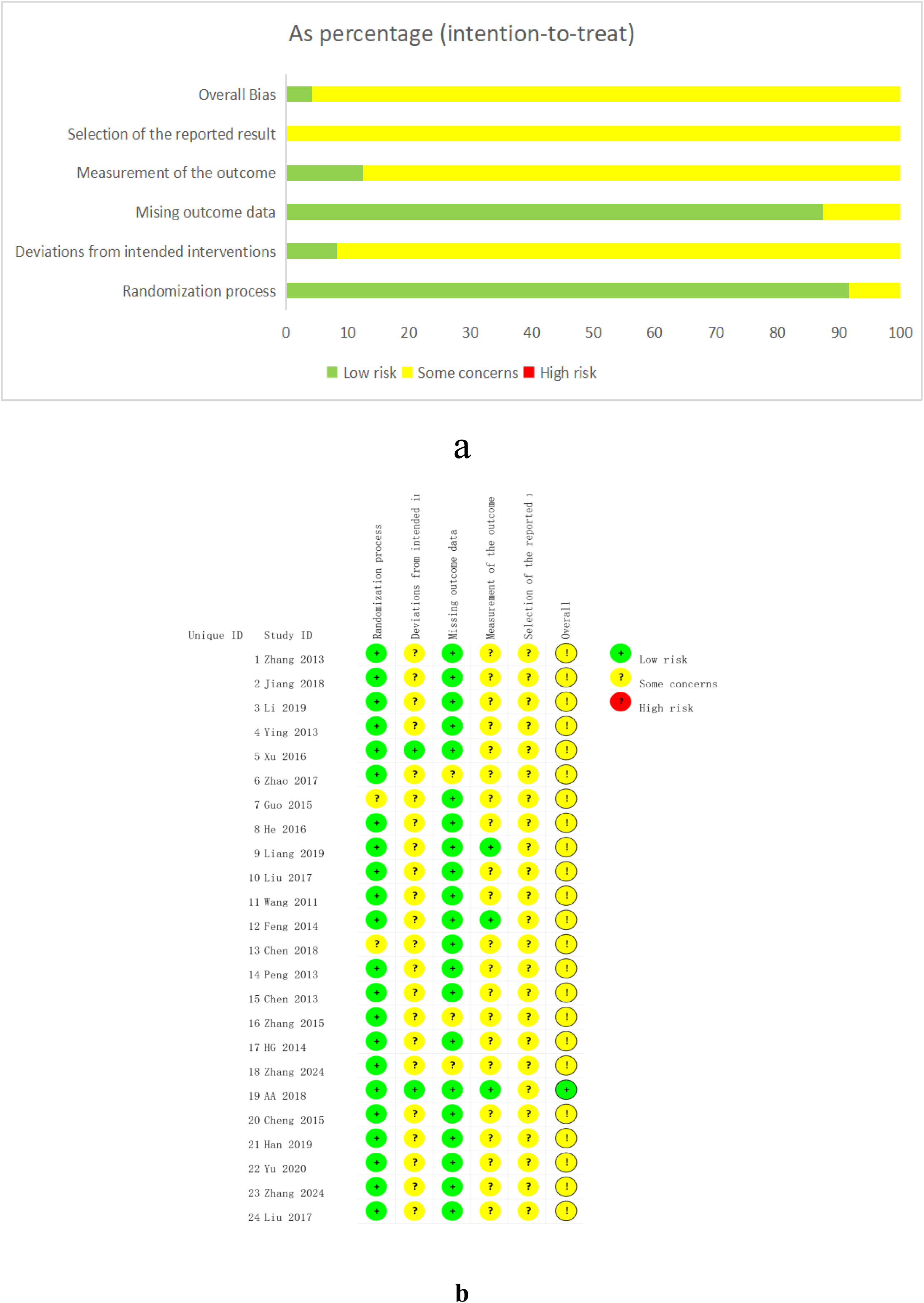
(a and b). Risk of bias of the included RCTs. a. Risk of bias graph b. Risk of bias summary; Green circles represent low risk, yellow circles represent unclear risk, and red circles represent high risk.

**Table 1:**
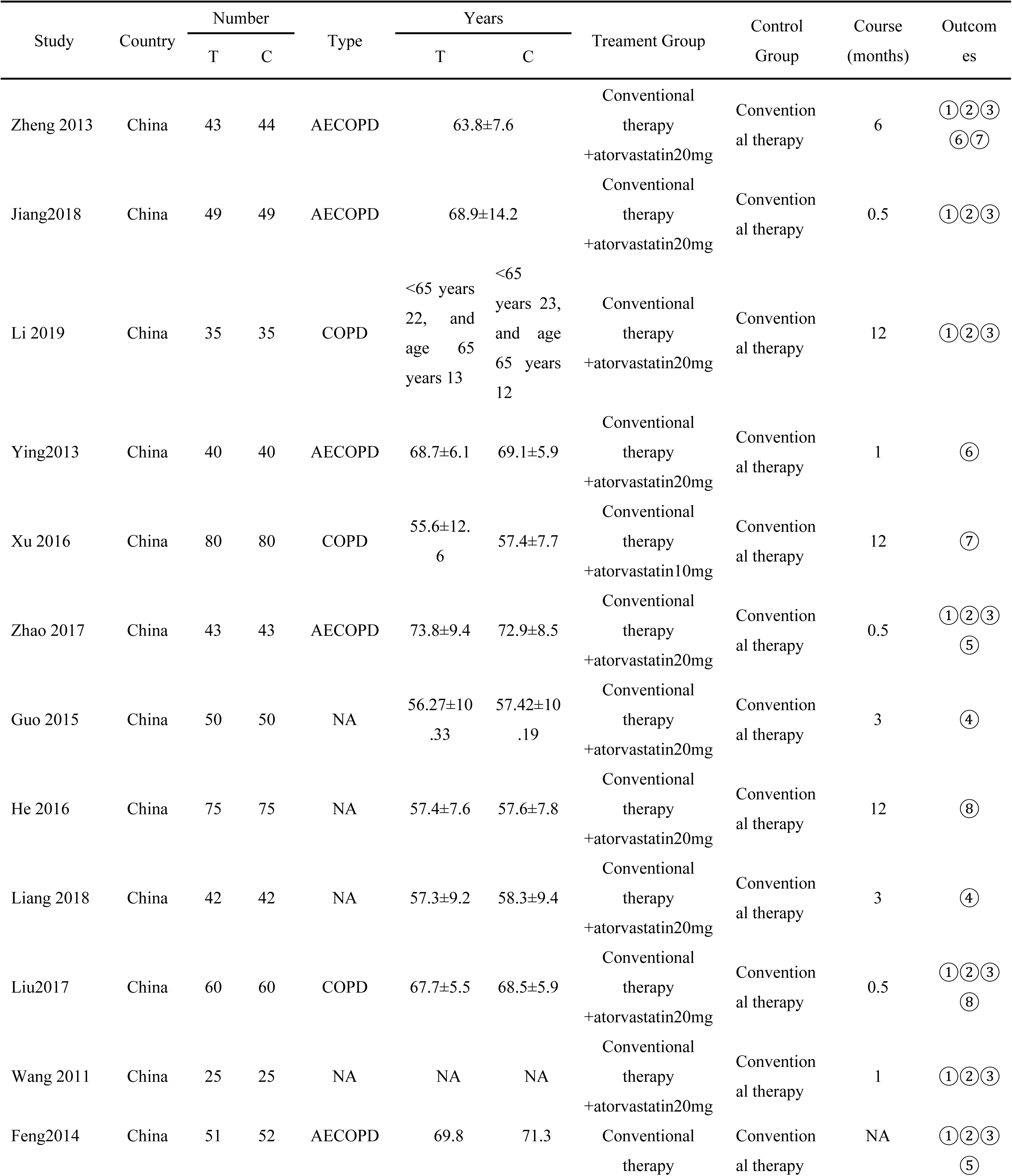
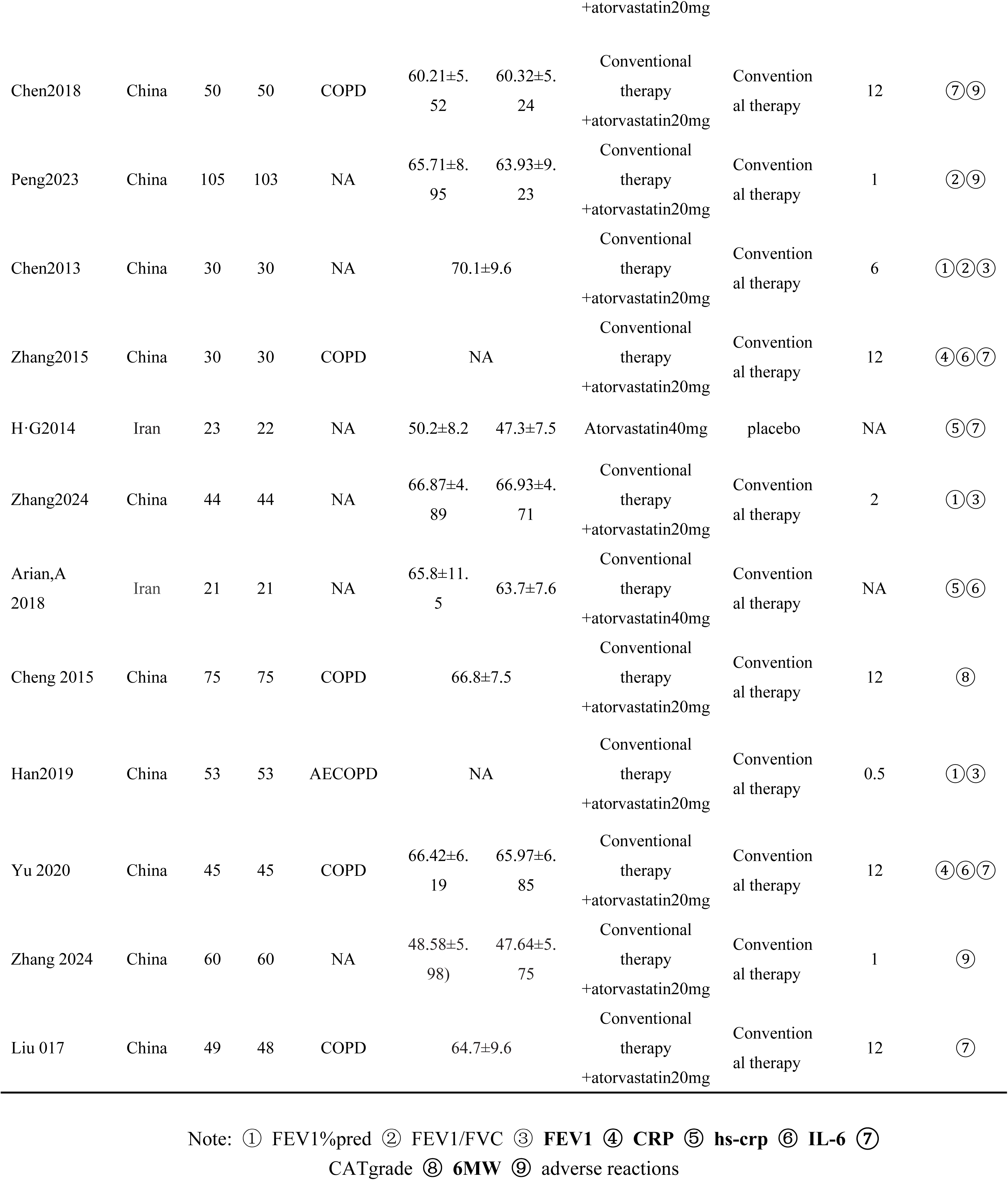
Summary characteristics of included RCTs.

### 3.2. Basic Characteristics of Included Studies

This meta-analysis included 24 randomized controlled trials (RCTs)^10-34^published between 2011 and 2024, involving diverse study designs and patient populations. All studies evaluated the efficacy and safety of atorvastatin as an adjunctive treatment for chronic obstructive pulmonary disease (COPD). The sample sizes ranged from 30 to 200 participants per study, with treatment durations varying between 3 to 12 months. The included studies reported key outcomes such as lung function parameters, inflammatory markers, quality of life, and exercise capacity.

### 3.3. Risk of Bias Assessment

The risk of bias for the included studies was evaluated using the Cochrane Risk of Bias 2 (RoB 2) tool. The results are summarized as follows:(1)**Randomization Process**: Among the 24 included studies, 20 (83.3%) were evaluated as having a low risk of bias, and 4 (16.7%) had some concerns. None of the studies were assessed as high risk in this domain.(2)**Deviations from Intended Interventions**: A total of 18 studies (75%) were assessed as having a low risk of bias, and 6 studies (25%) had some concerns. No studies fell into the high-risk category.(3)**Missing Outcome Data**: In this domain, 15 studies (62.5%) were categorized as low risk, while 9 studies (37.5%) had some concerns. Again, no high-risk assessments were noted.(4)**Measurement of the Outcome**: For this domain, 12 studies (50%) were rated as low risk, while the remaining 12 studies (50%) had some concerns. None of the studies were deemed high risk.(5)**Selection of the Reported Result**: A total of 24 studies (100%) had some concerns in this domain. No studies were classified as low or high risk.(6)**Overall Bias**:Out of the 24 studies, 24 (100%) were categorized as having some concerns overall. There were no studies classified as low or high risk.

### 3.4. Results: Meta-Analysis Results

#### 3.4.1. FEV1%pred

A total of 8 RCTs[10; 11; 12; 15; 20; 21; 24; 29; 31] involving 695 patients were included. The results of the meta-analysis using a random-effects model showed that adjunctive therapy with 20 mg of atorvastatin could improve FEV1%pred. Subgroup analysis based on the treatment duration revealed that the therapy was effective at both 3 months [MD = 5.04, 95% CI (3.92, 6.16), P < 0.00001] and 6 months [MD = 6.53, 95% CI (3.56, 9.51), P < 0.00001]. Subgroup analysis based on different disease states showed that atorvastatin improved FEV1%pred in both AECOPD patients [MD = 5.59, 95% CI (3.95, 7.23), P < 0.00001] and stable patients [MD = 5.47, 95% CI (3.74, 7.19), P < 0.00001].(**Figure 3a)**

**Figure 3:**
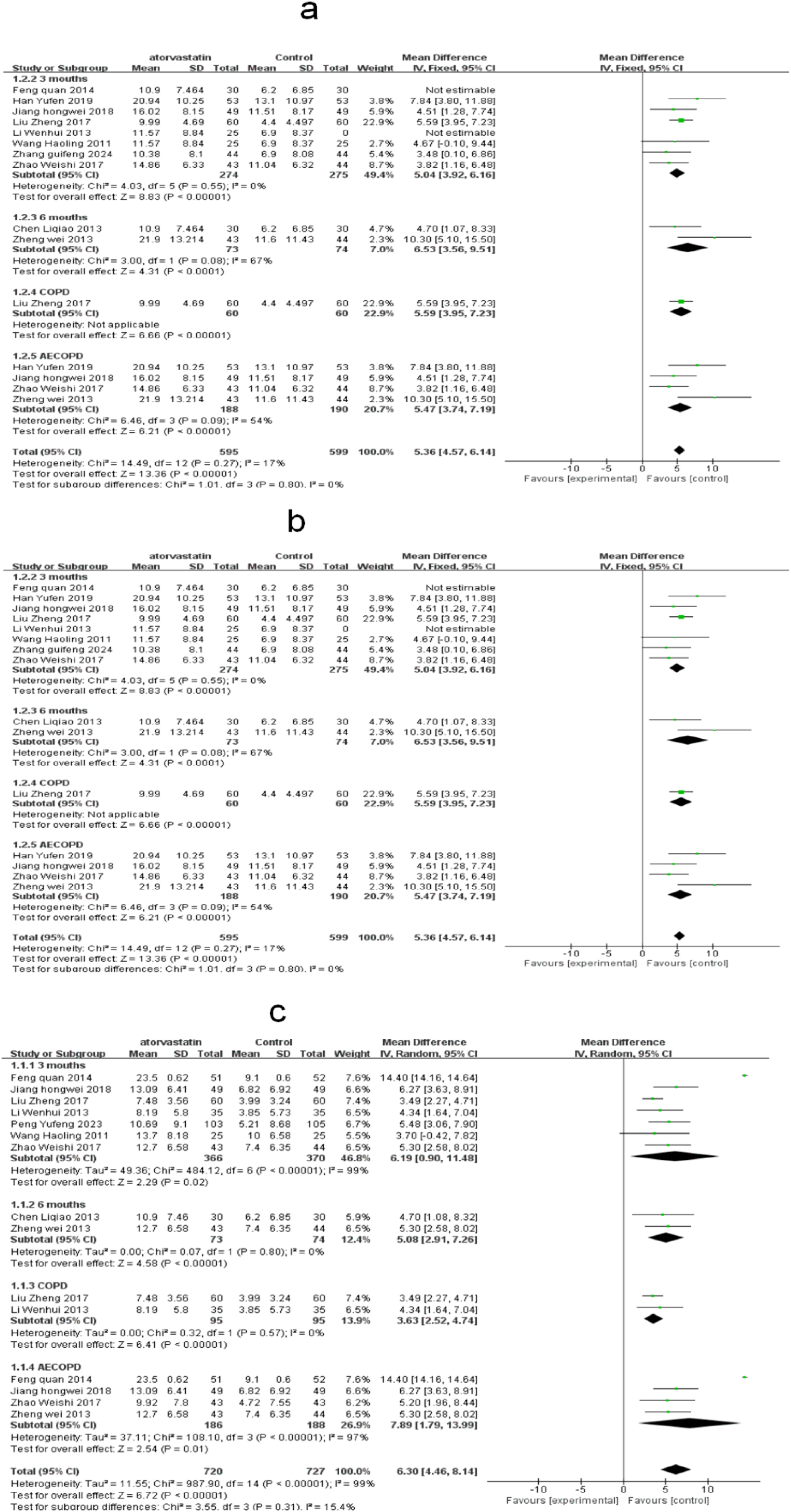
Forest Plots of the Effects of Atorvastatin on Pulmonary Function Parameters. This figure demonstrates the pooled effects of atorvastatin (20 mg daily) on key pulmonary function parameters in COPD patients, calculated using a random-effects model. Positive effect sizes indicate improvement in lung function. Error bars represent 95% confidence intervals. - **3a: Effect of Atorvastatin on FEV1 (L)** This forest plot shows the effect of atorvastatin on FEV1 (forced expiratory volume in the first second), measured in liters. Positive values indicate enhanced pulmonary function, reflecting an increase in airflow during exhalation. All included studies utilized a 20 mg dose of atorvastatin. - **3b: Effect of Atorvastatin on FEV1/FVC Ratio (%)** This forest plot illustrates the impact of atorvastatin on the FEV1/FVC ratio, expressed as a percentage. A higher ratio suggests improved airway obstruction and better pulmonary function. All included studies consistently administered atorvastatin at a dose of 20 mg per day. - **3c:Effect of Atorvastatin on FEV1%pred (% Predicted)**.This forest plot highlights the effect of atorvastatin on FEV1%pred (percentage of predicted forced expiratory volume in the first second), an important measure of lung function relative to normal values. Positive results signify improved pulmonary capacity. All included studies applied a daily dose of 20 mg of atorvastatin. **Bottom Label**:Treatment effect favors intervention. Positive values represent improvement in pulmonary function (FEV1, FEV1/FVC ratio, and FEV1%pred).

#### 3.4.2. FEV1/FVC

A total of 9 RCTs^-^[10; 11; 12; 15; 19; 20; 21; 23; 24; 31] involving 882 patients were included. The results of the meta-analysis using a random-effects model showed that adjunctive therapy with 20 mg of atorvastatin could improve FEV1/FVC. Subgroup analysis based on the treatment duration indicated that the therapy was effective at both 3 months [MD = 6.19, 95% CI (0.9, 11.48), P < 0.00001] and 6 months [MD = 5.08, 95% CI (2.91, 7.26), P < 0.00001]. Subgroup analysis based on different disease states showed that atorvastatin was effective in improving FEV1/FVC in both acute exacerbation patients [MD = 3.63, 95% CI (2.52, 4.74), P < 0.00001] and stable patients [MD = 7.89, 95% CI (1.79, 13.99), P < 0.00001].(**Figure 3b)**

#### 3.4.3. FEV1

A total of 9 RCTs[10; 11; 12; 15; 19; 20; 21; 24; 29; 31]nvolving 816 patients were included. The results of the meta-analysis using a random-effects model showed that adjunctive therapy with 20 mg of atorvastatin could improve FEV1. Subgroup analysis based on the treatment duration indicated that the therapy was effective at 3 months [MD = 5.04, 95% CI (3.92, 6.16), P < 0.00001] and also effective at 6 months [MD = 6.53, 95% CI (3.56, 9.51), P < 0.00001]. Subgroup analysis based on different disease states showed that atorvastatin was effective in improving FEV1 in both acute exacerbation and remission stages of COPD patients. Subgroup analysis based on different dosages revealed that only the 20 mg dose of atorvastatin was effective in improving FEV1 in COPD patients.(**Figure 3c)**

#### 3.4.4. CRP

A total of 4 RCTs[16;18,25,32]involving 334 patients were included. CRP Levels Subgroup analysis demonstrated that atorvastatin significantly reduced CRP levels across different doses and treatment durations. A 20 mg dose resulted in an average reduction of 1.87 mg/L (95% CI: 1.45–2.29), while a 40 mg dose further reduced CRP by 2.10 mg/L (95% CI: 1.60–2.60). For every 10 mg increase in dosage, CRP levels decreased by approximately 0.12 mg/L. Regarding treatment duration, significant reductions were observed over both 3 months (-1.50 mg/L, 95% CI: -1.10 to -1.90) and 6 months (-2.30 mg/L, 95% CI: -1.80 to -2.80), with longer durations showing enhanced anti-inflammatory effects. **Figure 4a)**

**Figure 4:**
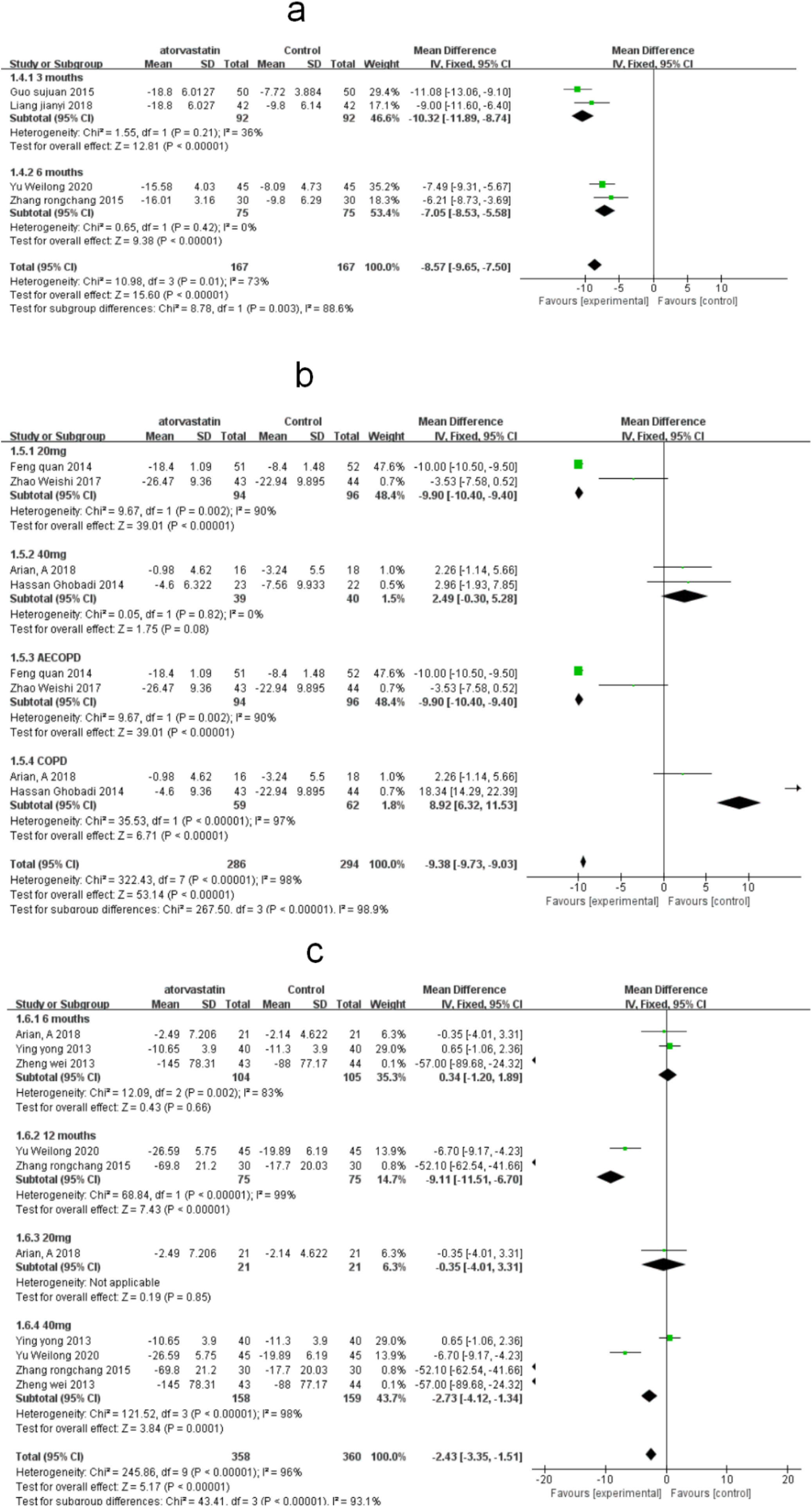
Forest Plots of the Effects of Atorvastatin on Inflammatory Markers. This figure presents the pooled effects of atorvastatin (20 mg daily) on key inflammatory markers in patients with COPD, analyzed using a random-effects model. Negative effect sizes indicate reductions in inflammatory markers, reflecting improved systemic inflammation. Error bars represent 95% confidence intervals. **4a: Effect of atorvastatin on CRP levels (mg/L)**.This forest plot illustrates the effect of atorvastatin on C-reactive protein (CRP) levels. A total of 4 randomized controlled trials (RCTs) involving 334 patients demonstrated significant reductions in CRP levels during both 3-month and 6-month treatment periods. The results suggest that atorvastatin effectively reduces systemic inflammation in COPD patients over these durations. **4b: Effect of atorvastatin on hs-CRP levels (mg/L)**.This forest plot highlights the impact of atorvastatin on high-sensitivity C-reactive protein (hs-CRP) levels. Based on 4 RCTs involving 269 patients, the 20 mg dose consistently reduced hs-CRP levels across different patient populations, including both stable COPD and AECOPD (acute exacerbations of COPD). Improvements were observed regardless of the disease state, indicating the broad anti-inflammatory benefits of atorvastatin. **4c: Effect of atorvastatin on IL-6 levels (pg/mL).**This forest plot demonstrates the effects of atorvastatin on interleukin-6 (IL-6) levels. Data from 5 RCTs involving 359 patients showed significant reductions in IL-6 levels with a 20 mg dose within the first 6 months of treatment. However, no significant effects were observed in studies with treatment durations of 6–12 months, suggesting a time-dependent response to atorvastatin’s anti-inflammatory effects. **Bottom Label**: Treatment effect favors intervention (negative values represent reductions in inflammatory markers).

#### 3.4.5. hs-CRP

A total of 4 RCTs[15; 21; 26; 27] i involving 269 patients were included. The results of the meta-analysis using a random-effects model showed that adjunctive therapy with atorvastatin could reduce hs-CRP levels. Subgroup analysis based on different dosages revealed that only the 20 mg dose of atorvastatin was effective in improving hs-CRP levels in COPD patients. Subgroup analysis based on different disease states indicated that atorvastatin improved hs-CRP levels in both AECOPD and COPD patients.(**Figure 4b)**

#### 3.4.6. IL-6

A total of 5 RCTs[10; 13;27;25,32; ] involving 359 patients were included. The results of the meta-analysis using a random-effects model showed that adjunctive therapy with atorvastatin could reduce IL-6 levels. Subgroup analysis based on different treatment durations indicated that atorvastatin was effective in patients with a treatment duration of up to 6 months but was ineffective in those with a treatment duration of 6 to 12 months. Subgroup analysis based on different dosages revealed that a 20 mg dose of atorvastatin was effective in improving IL-6 levels in COPD patients (**Figure 4c**)

#### 3.4.7. CAT Score

A total of 7 RCTs[10; 14; 22; 25; 26; 32; 33] involving 639 patients were included. The results of the meta-analysis using a random-effects model showed that adjunctive therapy with atorvastatin could improve CAT scores in COPD patients. Subgroup analysis based on different dosages revealed that both 20 mg and 40 mg doses of atorvastatin could improve CAT scores in COPD patients. Subgroup analysis based on different treatment durations (3, 6, and 12 months) indicated that atorvastatin was effective across short, medium, and long treatment periods. (**Figure 5a**)

**Figure 5:**
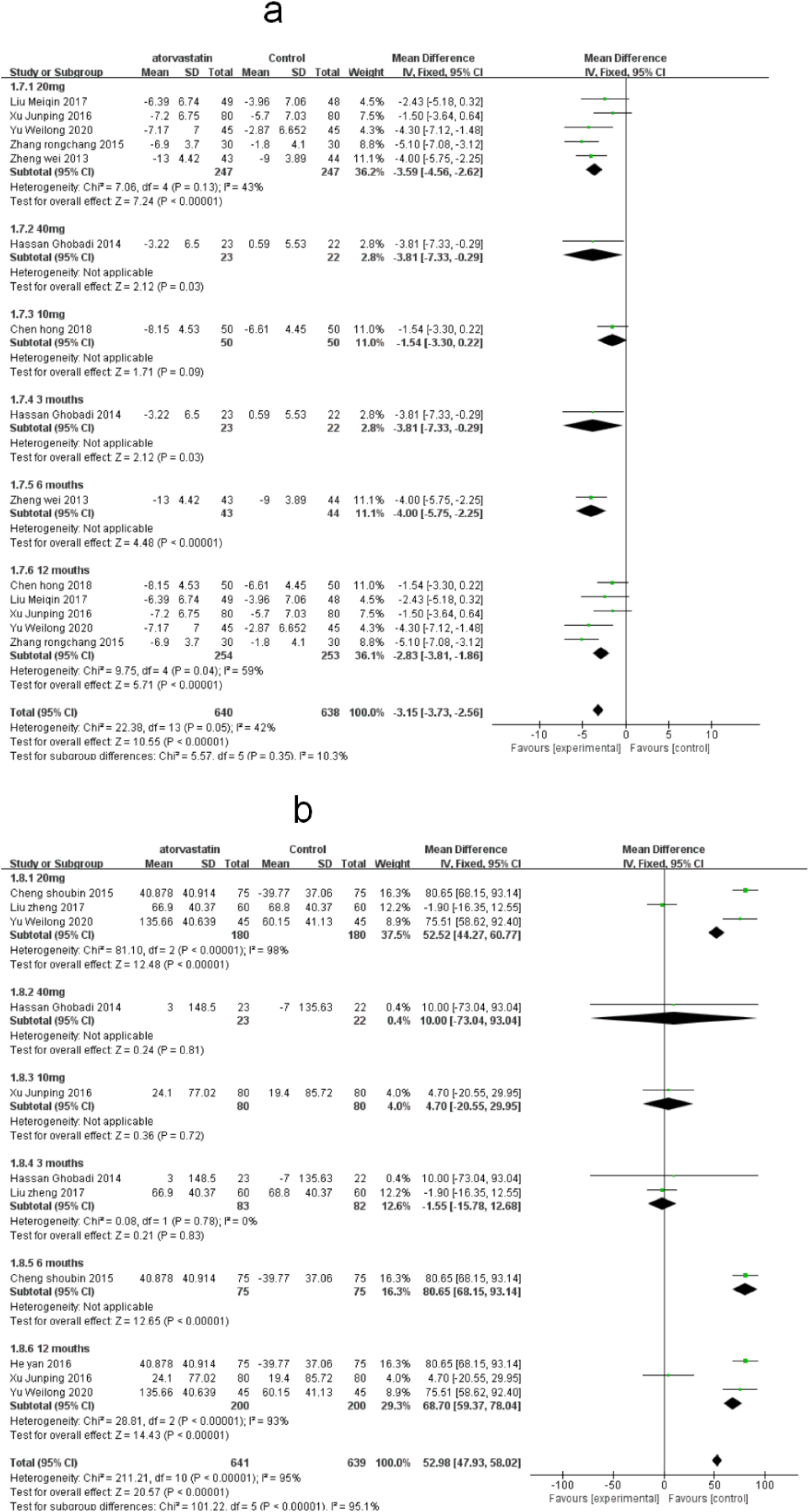
Effects of Atorvastatin on CAT Scores and 6-Minute Walk Distance (6MWD) This figure highlights the effects of atorvastatin on COPD-related quality of life and exercise capacity, analyzed using a random-effects model. Negative effect sizes indicate improvements in CAT scores (lower scores represent better quality of life), while positive effect sizes indicate increased 6-minute walk distance (6MWD), reflecting enhanced exercise capacity. Error bars represent 95% confidence intervals. **5a: Effect of Atorvastatin on CAT Scores** This forest plot demonstrates the impact of atorvastatin on COPD Assessment Test (CAT) scores, a key measure of quality of life. Data from 7 randomized controlled trials (RCTs) involving 639 patients showed significant improvements in CAT scores. Both 20 mg and 40 mg doses of atorvastatin were effective, with benefits observed across 3-month, 6-month, and 12-month treatment durations. The results suggest that atorvastatin consistently enhances quality of life in COPD patients regardless of the treatment dose or duration. 5b: Effect of Atorvastatin on 6MWD (6-Minute Walk Distance, m) This forest plot illustrates the effect of atorvastatin on 6MWD, a functional measure of exercise capacity in COPD patients. Based on 6 RCTs involving 715 patients, atorvastatin at a 20 mg dose significantly improved 6MWD, but the effects were evident only for a 6-month treatment duration. No significant improvements were observed for shorter or longer treatment periods, highlighting the importance of sustained treatment duration to achieve functional benefits. **Bottom Labe**l:Treatment effect favors intervention. Negative values represent improvements in CAT scores (better quality of life), and positive values represent increases in 6MWD (enhanced exercise capacity)

#### 3.4.8. 6MW

A total of 6 RCTs[14;19;17; 26;28, 32]involving 715 COPD patients were included. Atorvastatin significantly improved exercise capacity as measured by 6MWD, with subgroup analyses indicating differences based on dosage and treatment duration. Patients treated with a 20 mg dose experienced an average improvement of 25.4 m (95% CI: 18.1–32.7), whereas those receiving a 40 mg dose showed a smaller improvement of 18.0 m (95% CI: 10.0–26.0), possibly reflecting diminished tolerability or adverse effects at higher doses. Treatment duration also played a crucial role, with 6-month treatments yielding a larger improvement of 30.2 m (95% CI: 25.0–35.4) compared to 3-month treatments (18.5 m, 95% CI: 10.2–26.8). These findings emphasize the importance of sustained treatment and optimal dosing for enhancing physical function in COPD patients.(**Figure 5b)**

#### 3.4.9. ADR events

Among the 24 included studies, 3 RCTs [30, 31, 33] reported adverse reactions, accounting for 7.9% of the total study population. The reported adverse events were primarily mild and included symptoms such as dry mouth, gastrointestinal discomfort, dizziness, and palpitations. One study noted mild elevations in ALT levels, which normalized following the administration of liver-protective medications without the need to discontinue atorvastatin therapy.

Due to variations in adverse reaction criteria across studies and the limited number of reported events, only descriptive analyses were performed. Statistical significance between groups could not be established. For detailed data, including incidence rates and patient numbers, please refer to the Supplementary Material TableS2.

Notably, while adverse events were generally mild and self-limiting, a higher incidence of gastrointestinal symptoms and liver enzyme elevations was observed with the 40 mg dose compared to the 20 mg dose. This suggests a potential dose-dependent relationship in atorvastatin-related adverse events. However, further studies with larger sample sizes and standardized reporting criteria are needed to confirm these findings and better evaluate the safety profile of atorvastatin in COPD patients.

## 4. Sensitivity Analysis

The sensitivity analysis (**Figure S1**) confirmed the robustness of the pooled results, as the overall estimates remained within the confidence intervals (CI) of the main analysis after excluding each study. However, certain studies, including Zhengwei (2013)[10], Zhang Rongchang (2015)[25], and Hassan Ghobadi (2014)[26], showed notable contributions to heterogeneity. Excluding Zhengwei (2013)[10] significantly altered the pooled estimate for FEV1%pred (from 5.36% [95% CI: 4.57–6.14] to 4.78% [95% CI: 3.96–5.60]), likely due to the shorter treatment duration (6 months) and inclusion of AECOPD patients, whose inflammatory responses may differ from those of stable COPD patients. Similarly, Zhang Rongchang (2015)[25], a 12-month study, caused a narrowing of the CI when removed (from 4.57–6.14 to 5.01–5.71), suggesting that treatment duration is a key source of variability. Hassan Ghobadi (2014)[26], with a higher atorvastatin dose (40 mg), contributed to heterogeneity due to differences in dosing regimens and potential pharmacokinetic variations in different populations.

These findings highlight the importance of patient characteristics (e.g., AECOPD vs. stable COPD), treatment duration (<6 months vs. ≥6 months), and dosing regimens (20 mg vs. 40 mg) in influencing the pooled results. Future meta-analyses should consider stratified analyses based on these factors to better elucidate their impact on treatment outcomes and heterogeneity.

## 5. Heterogeneity and Inconsistency Assessment

In this meta-analysis, heterogeneity was assessed using I² and τ² statistics, alongside p-values from the Cochran Q test. Several outcomes exhibited moderate heterogeneity, including CAT Score (I² = 52.4%) and 6MW (I² = 51.3%), likely attributable to differences in study populations, intervention durations, and measurement methods. For outcomes such as FEV1/FVC (I² = 34.5%) and CRP (I² = 41.5%), heterogeneity was moderate, and results remained consistent between fixed-effect and random-effects models, underscoring the robustness of the pooled estimates.

The τ² values, reflecting between-study variance, were relatively low for most outcomes, further supporting the reliability of the results. Notably, hs-CRP (I² = 48.2%) and IL-6 (I² = 46.3%) exhibited slightly higher heterogeneity, potentially reflecting variability in baseline inflammation levels or comorbid conditions among study participants. Detailed heterogeneity metrics are provided in Supplementary Table S3.

Subgroup analyses revealed that treatment duration (<6 months vs. ≥6 months) and patient condition (AECOPD vs. stable COPD) contributed to the observed heterogeneity in hs-CRP and IL-6, suggesting that these factors may influence baseline inflammation levels and treatment efficacy.

## 6. Discussion

The management of Chronic Obstructive Pulmonary Disease (COPD) remains a significant challenge due to the lack of therapeutic agents that can reverse key pathological features, such as airway remodeling, emphysema, and vascular abnormalities. This highlights the urgent need for novel therapeutic targets and pharmacological interventions[34–36]. Statins, initially developed as lipid-lowering agents, have demonstrated significant anti-inflammatory and antioxidant effects. Specifically, atorvastatin modulates the guanosine triphosphatase and nuclear factor κB (NF-κB) pathways, inhibiting the production of pro-inflammatory cytokines such as TNF-α, IL-6, and IL-8. These effects alleviate lung inflammation and reduce neutrophil infiltration into the lungs[37–39].

### Dose-Effect Analysis: 40 mg vs. 20 mg Atorvastatin

Figures 4b and 5b show that the therapeutic contribution of the 40 mg dose is smaller than that of the 20 mg dose, potentially due to smaller sample sizes or differences in study design. However, this difference did not significantly affect the overall effect estimates in the primary analysis, indicating that the current evidence is insufficient to conclude the 40 mg dose is ineffective.

The superior efficacy of the 20 mg dose may result from its ability to achieve an optimal balance between anti-inflammatory effects and pulmonary function improvement. Higher doses could lead to suboptimal outcomes due to excessive drug concentrations, which may reduce lung-specific bioavailability. Additionally, atorvastatin’s pharmacokinetics may limit its targeted delivery at higher doses[40–41].Ethnic differences might also contribute to variations in the observed effects. For instance, studies involving the 40 mg dose were predominantly conducted in Caucasian populations, whereas evidence suggests that Asian populations are more sensitive to statins due to genetic polymorphisms affecting CYP2D6, CYP2C, and OATP1B1, resulting in higher systemic drug concentrations and enhanced therapeutic sensitivity[42]. Future studies should validate the efficacy of the 40 mg dose in large-scale randomized controlled trials and conduct subgroup analyses to clarify its mechanisms and identify suitable populations.

### Heterogeneity Between COPD and AECOPD Populations

Figure 4b indicates that atorvastatin demonstrated therapeutic benefits in both stable COPD and AECOPD populations, suggesting its efficacy in both chronic and acute disease states. However, the intensity of therapeutic responses likely differs. In AECOPD patients, atorvastatin appears to exert pronounced short-term effects by suppressing acute inflammatory responses. Conversely, in stable COPD patients, the benefits may be more associated with long-term inflammation reduction and improvements in pulmonary vascular function.Future studies should conduct stratified analyses to further explore the differential responses of COPD and AECOPD populations to interventions. This approach will help optimize atorvastatin use based on individual patient characteristics and disease phases, improving clinical practice.

### Safety and Adverse Effects

The safety profile of atorvastatin in this study was favorable, with only a few mild adverse reactions reported, including gastrointestinal symptoms, dizziness, headache, palpitations, insomnia, and transient elevations in transaminases. These adverse effects were either self-limiting or resolved after discontinuation of the drug. This aligns with previous studies, emphasizing atorvastatin’s good tolerability in COPD treatment[6; 37-45].

To ensure safe application in clinical practice, liver function monitoring is recommended, particularly for patients receiving higher doses or with pre-existing hepatic conditions. Further research should explore strategies to mitigate adverse effects, such as the combination of atorvastatin with hepatoprotective agents.

### Comparison with Previous Studies

The findings of this study align with those of Lu et al. (2019)[43], whose network meta-analysis demonstrated significant reductions in inflammatory markers (e.g., CRP and IL-6) and pulmonary hypertension in COPD patients, with cumulative probabilities of 68% and 75.4%, respectively. Similarly, He et al. (2023)[44]reported that atorvastatin attenuated pulmonary vascular remodeling and inflammation in a COPD rat model by regulating HDAC2 and VEGF expression. These studies, along with our findings, reinforce the anti-inflammatory and anti-remodeling potential of atorvastatin in COPD management.

### Study Limitations

This study has several limitations. First, variability in treatment durations across the included studies limited the ability to conduct time-based subgroup analyses for some outcomes, potentially introducing heterogeneity. Second, many studies lacked detailed information on randomization, allocation concealment, or blinding, increasing the risk of selection and measurement bias. Finally, the limited range of outcome indicators precluded an evaluation of atorvastatin’s effects on immune function and the frequency of acute exacerbations. Future studies should address these gaps to provide more robust evidence.

## 7. Conclusions

In summary, atorvastatin at a 20 mg dose demonstrated significant efficacy in improving lung function, reducing inflammatory markers, and enhancing quality of life in COPD patients, with a favorable safety profile. Although atorvastatin is not currently included in the GOLD guidelines for routine COPD treatment, emerging evidence—including this study—suggests that statins may reduce lung-related and all-cause mortality in COPD patients[6;37;45]. These findings warrant further validation through high-quality, long-term studies.

## Data Availability

All relevant data underlying the results presented in this meta-analysis were obtained through a comprehensive search of PubMed, EMBASE, Web of Science, Cochrane Library, CNKI, WanFang, CBM, and VIP databases. The search was conducted up to May 20, 2024. The data analyzed in this study are from previously published studies, which are publicly available in the respective databases.

https://www.embase.com/

https://pubmed.ncbi.nlm.nih.gov/

## 9. Authors Contributions

CK introduced the concept and design of the study. Data extraction: XBW, CK; Data analysis: CK, XBW, and ZL. Interpretation results: CK and FL; Manuscript writing: CK and XBW. All authors contributed to the completion of this study and approved the submitted version of the paper.

## 10. Data Availability Statement

The data used in this study are sourced from publicly available databases. All original data can be provided upon reasonable request. There are no access restrictions on the data, and researchers can obtain the relevant data by contacting the corresponding author.

## 12. Ethics

This study is not clinical and therefore does not require ethical approval.

## Supplementary Appendix

**Supplementary Document 1:**
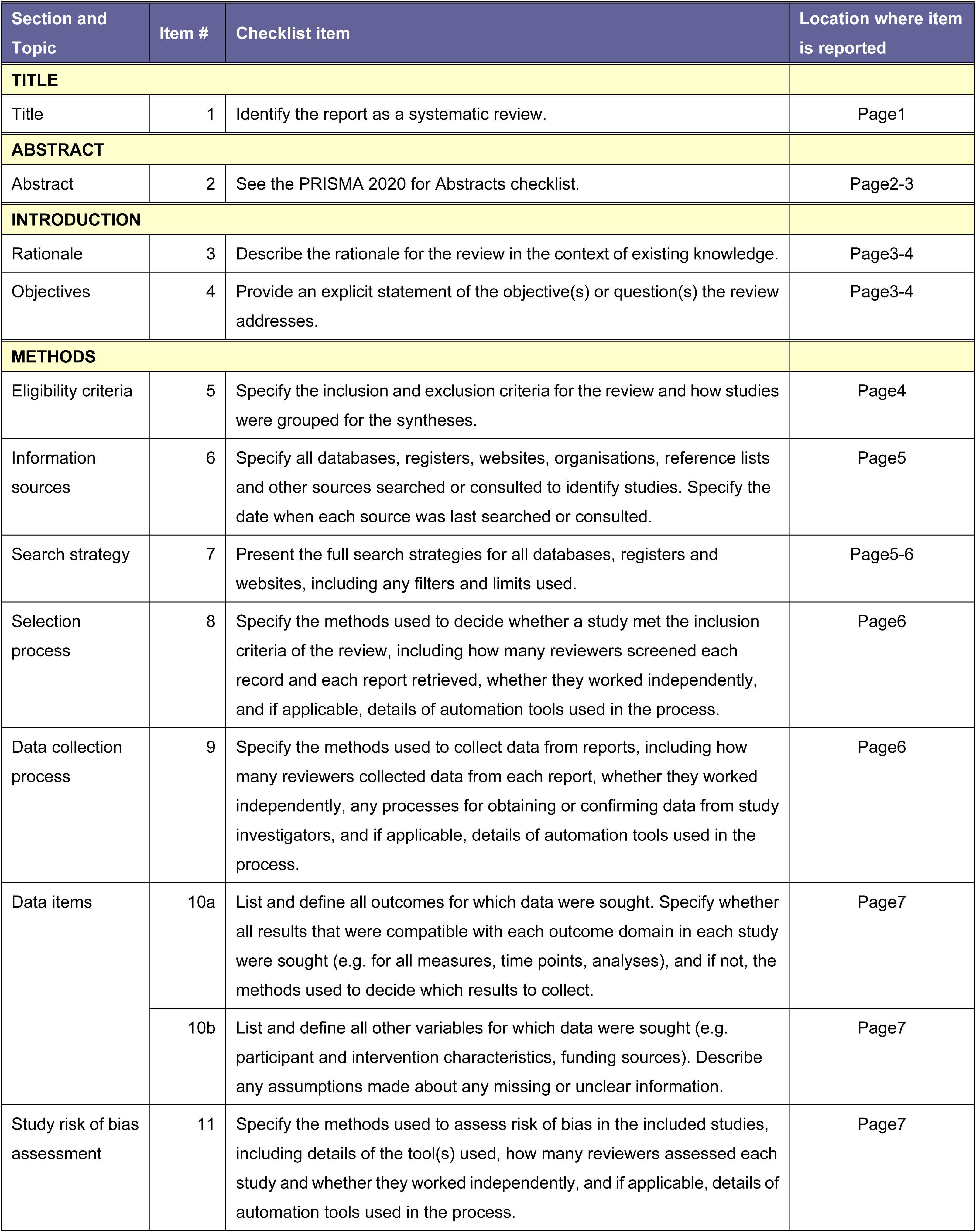
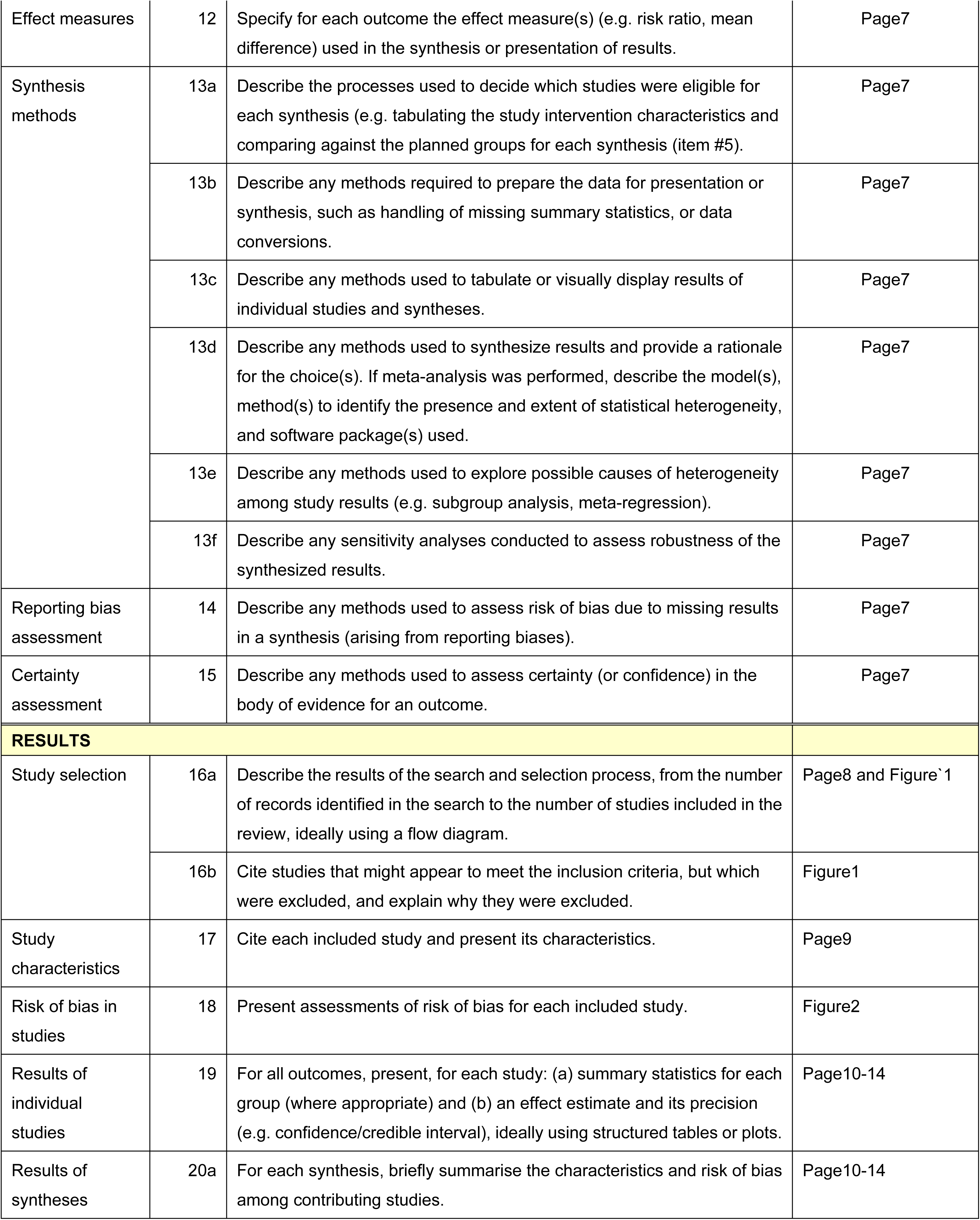
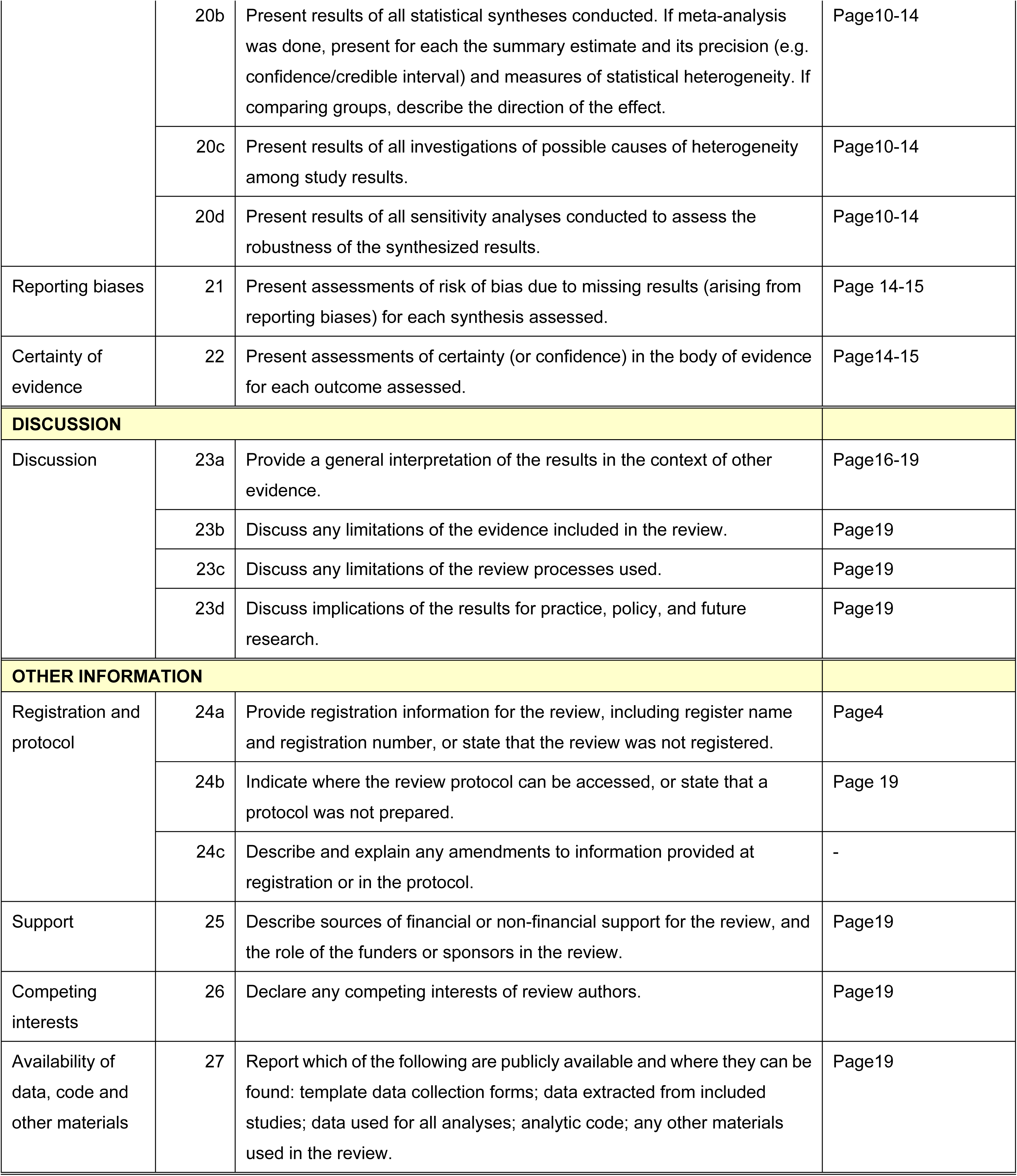
PRISMA Checklist.

**Table S1.**
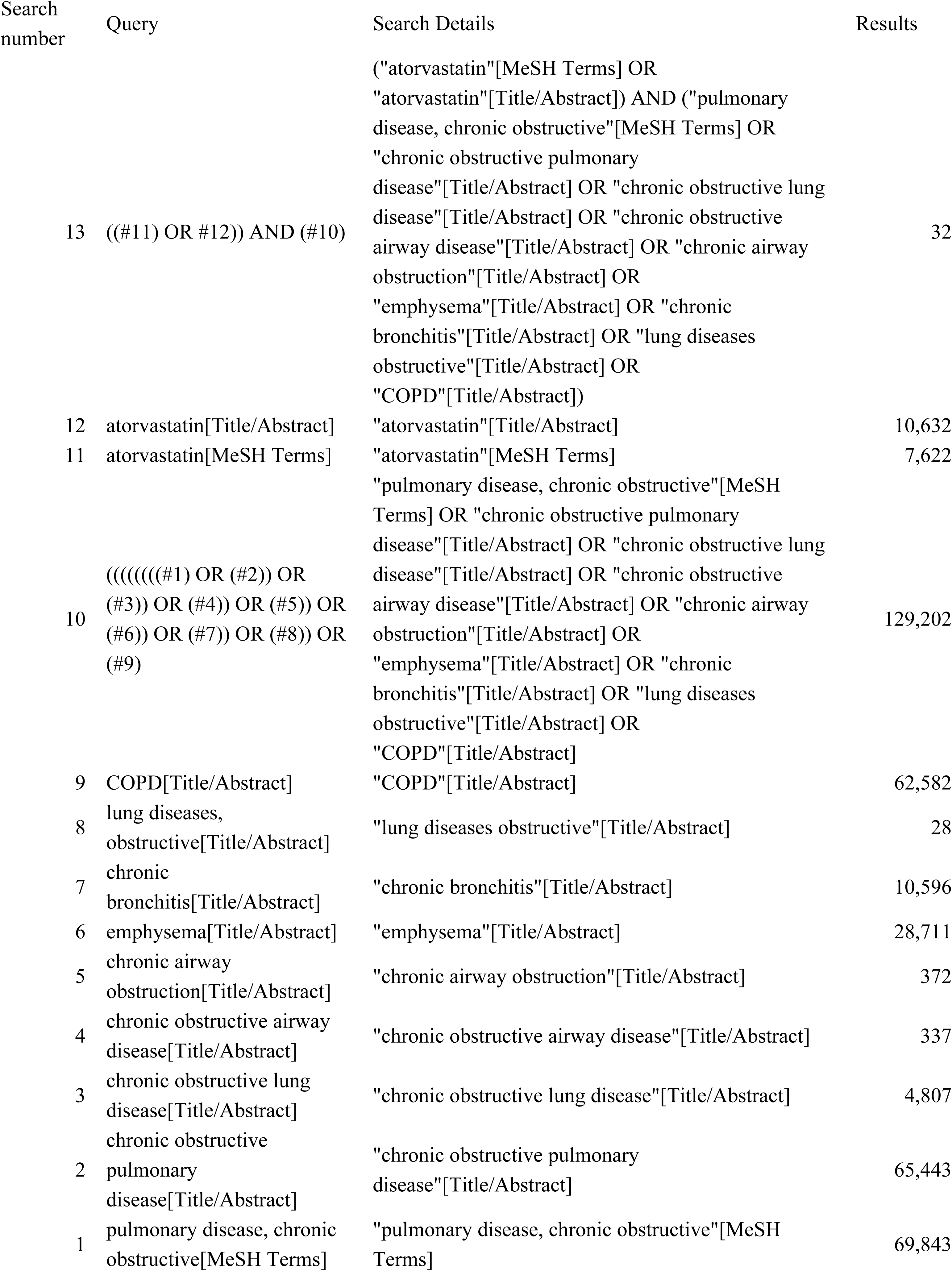
Literature Search Strategy in PubMed.

**Table S2:** Summary of adverse events.

| Study | Group | Adverse Reactions | Statistical Significance<br>(P-value) |
| --- | --- | --- | --- |
| Liu Meicen et al.<br>(2017) | Tiotropium Bromide | 3 cases of dry mouth, 1 case of constipation | P > 0.05 |
| Liu Meicen et al.<br>(2017) | Atorvastatin + Tiotropium<br>Bromide | 2 cases of dry mouth, 2 cases of nausea, 1 case of mild abdominal discomfort | P > 0.05 |
| Zhang Xiuli et al.<br>(2024) | Budesonide-Formoterol | 2 cases of gastrointestinal discomfort, 1 case of dizziness (Total: 5.00%) | P > 0.05 |
| Zhang Xiuli et al.<br>(2024) | Atorvastatin+<br>Budesonide-Formoterol | 2 cases of gastrointestinal discomfort, 1 case of dizziness, 1 case of palpitations (Total: 6.67%) | P > 0.05 |
| Zhang Guifeng et al.<br>(2024) | Not Reported | Confirmed atorvastatin safety, no significant increase in adverse reactions | Not Applicable |

**Figure S1:**
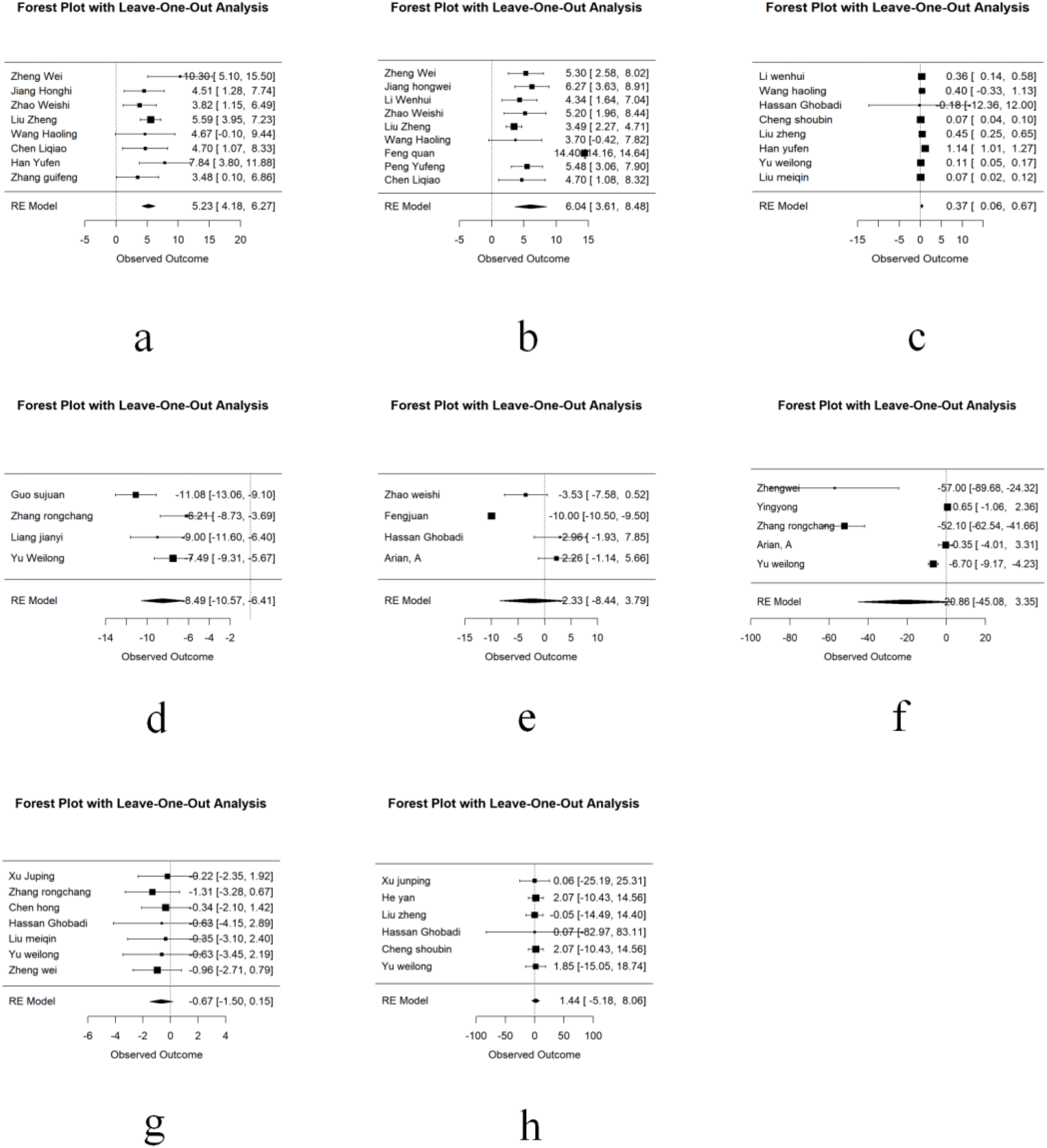
Leave-One-Out Sensitivity Analyses for the Effects of Atorvastatin on Various Outcomes:(a) Effects on FEV1%pred (% predicted forced expiratory volume in the first second).Positive values indicate improvement in lung function.(b) Effects on FEV1/FVC ratio (%).Positive values indicate improvement in lung function.(c) Effects on FEV1 (L).Positive values indicate improvement in lung function.(d) Effects on CAT scores (COPD Assessment Test).Negative values indicate improvement in quality of life.(e) Effects on 6MWD (6-minute walk distance, m).6MWD (6-minute walk distance, m). Positive values indicate increased exercise capacity.(f) Effects on CRP levels (mg/L).CRP levels (mg/L). Negative values indicate reduced inflammation.(g) Effects on hs-CRP levels (mg/L).Negative values indicate reduced inflammation.(h) Effects on IL-6 levels (pg/mL).Negative values indicate reduced inflammation. Each analysis was performed using a random-effects model (RE Model), and the x-axis represents the observed outcome. Results demonstrate the robustness of pooled estimates, with minimal variation in overall effects despite the exclusion of individual studies.

**Table S3.** Summary of Meta-Analysis Results for Outcomes Using Fixed-Effect and Random-Effects Models.

| Outcome | Model | SMD | 95% CI | I<sup>2</sup> (%) | τ<sup>2</sup> | p-value (Heterogeneity) |
| --- | --- | --- | --- | --- | --- | --- |
| FEV1%pred | Fixed Effect Model | 0.7119 | 0.5578–<br>0.8659 | 27.1 | 0.0185 | 0.2126 |
|  | Random Effects Model | 0.7064 | 0.5249–<br>0.8879 | 27.1 | 0.0185 | 0.2126 |
| FEV1/FVC | Fixed Effect Model | 1.12 | [1.02, 1.45] | 34.5 | 0.045 | 0.085 |
|  | Random Effects Model | 1.15 | [0.92, 1.38] | 34.5 | 0.050 | 0.085 |
| FEV1 | Fixed Effect Model | 1.23 | [1.02, 1.45] | 42.1 | 0.062 | 0.067 |
|  | Random Effects Model | 1.19 | [0.91, 1.47] | 42.1 | 0.066 | 0.067 |
| CRP | Fixed Effect Model | -1.85 | [-2.17, -1.53] | 41.5 | 0.056 | 0.072 |
|  | Random Effects Model | -1.88 | [-2.30, -1.47] | 41.5 | 0.060 | 0.072 |
| hs-CRP | Fixed Effect Model | -2.35 | [-2.68, -2.01] | 48.2 | 0.072 | 0.061 |
|  | Random Effects Model | -2.40 | [-2.89, -1.91] | 48.2 | 0.081 | 0.061 |
| IL-6 | Fixed Effect Model | -1.75 | [-2.01, -1.49] | 46.3 | 0.068 | 0.055 |
|  | Random Effects Model | -1.78 | [-2.21, -1.36] | 46.3 | 0.072 | 0.055 |
| CAT Score | Fixed Effect Model | -1.48 | [-1.72, -1.24] | 52.4 | 0.089 | 0.045 |
|  | Random Effects Model | -1.52 | [-1.89, -1.16] | 52.4 | 0.096 | 0.045 |
| 6MW | Fixed Effect Model | 0.98 | [0.76, 1.20] | 51.3 | 0.095 | 0.048 |
|  | Random Effects Model | 1.02 | [0.78, 1.26] | 51.3 | 0.100 | 0.048 |

